# Spinocerebellar ataxia type 4 is caused by a GGC expansion in the *ZFHX3* gene and is associated with prominent dysautonomia and motor neuron signs

**DOI:** 10.1101/2023.10.03.23296230

**Authors:** Martin Paucar, Daniel Nilsson, Martin Engvall, José Laffita-Mesa, Cilla Söderhäll, Mikael Skorpil, Christer Halldin, Patrik Fazio, Kristina Lagerstedt-Robinson, Göran Solders, Maria Angeria, Andrea Varrone, Mårten Risling, Hong Jiao, Inger Nennesmo, Anna Wedell, Per Svenningsson

## Abstract

**Background:** Spinocerebellar ataxias (SCAs) are a group of heterogeneous autosomal dominant disorders. Spinocerebellar ataxia 4 (SCA4), one of the rarest SCAs, is characterized by adult-onset ataxia, polyneuropathy and linked to chromosome 16q22.1, the underlying mutation remains to be discovered.

**Methods:** Three Swedish families affected by undiagnosed SCA went through detailed examinations, neurophysiological tests, neuroimaging studies and genetic testing. Imaging included MRI of the neuroaxis and brain PET. In 3 cases neuropathological assessments were performed. Genetic testing included SR WGS, STR analysis with Expansion Hunter *de novo* and long read (LR) WGS.

**Results:** Novel features such as dysautonomia, motor neuron affection, and abnormal eye movements, were found. Anticipation was documented as well. Atrophy in the cerebellum, brainstem and spinal cord was found in patients studied with MRI. [18F]FDG-PET demonstrated brain hypometabolism whereas [11C]Flumazenil-PET yielded reduced binding in several brain lobes, insula, thalamus, hypothalamus and cerebellum. Neuropathological assessment revealed moderate to severe loss of Purkinje cells in the cerebellum. In the spinal cord a marked loss of motor neurons in the anterior horns was seen as well as pronounced degeneration of posterior tracts. Intranuclear, mainly neuronal, inclusions positive for p62 and ubiquitin were sparse but widespread in the CNS. This finding prompted assessment for nucleotide expansions. A poly-glycine stretch encoding GGC expansions in the last exon of the zink finger homeobox 3 (*ZFHX3)* gene was identified segregating with disease, and not found in 1000 controls.

**Conclusions:** SCA4 is a neurodegenerative disease, caused by a novel GGC expansion in *ZFHX3,* arguably the first polyglycine disorder in humans.

## Introduction

Spinocerebellar ataxias (SCA) are a heterogeneous group of autosomal dominant (AD) diseases enumerated in order of disease discovery (1,2,3). Polyglutamine (polyQ) SCA represent up to 60% of all SCA cases, the remaining SCA subtypes are rare and display variable geographic distribution (1, 2). Spinocerebellar ataxia type 4 (SCA4) was initially described in a Scandinavian-American kindred from Utah/Wyoming and found to be linked to 16q22.1. Its phenotype consists of slowly progressive adult-onset ataxia and axonal sensory neuropathy (Flanigan et al 1996). Flanigan et al suggested that SCA4 was synonymous with Biemond’s ataxia (4, 5). A second SCA4 family was described in Germany with neuroimaging and neuropathological data provided for this disease (6, 7). Cerebellar atrophy was reported for SCA4, whereas the neuropathological assessment included the brainstem and cerebellum only (6, 7). Anticipation has been suggested in these SCA4 families which motivated a screening for CAG/CTG expansions in a narrowed candidate region but yielded negative results (6). In addition, staining with the anti-polyglutamine antibody 1C2 did not yield any reactivity (7). Here we report novel findings for SCA4 such as dysautonomia, motor neuron symptoms and signs, eye movement abnormalities, and atrophy of both the pons and spinal cord. For the first time for this disease, brain PET data is provided.

The candidate region 16q22.1 in SCA4 (4), was narrowed from 6 cM to a 3.69 cM interval (6). Here, we further narrowed the candidate region and provide a detailed characterization of three SCA4 kindreds in Sweden. The presence of intranuclear inclusions positive for p62 and ubiquitin scattered neurons across the central nervous system (CNS), typical for nucleotide expansions, oriented our search. Whole genome sequencing (WGS) and analysis with Expansion Hunter *de novo* (8) accordingly detected a heterozygous (GGC)_n_ nucleotide expansion in the final exon of the *ZFHX3* (*ATBF1*) gene, encoding a poly-glycine stretch in the ZFHX3 protein.

## Materials and methods

For further details on all material and methods, see supplementary material.

### Patients

The Swedish Ethical Review Authority (*Etikprövningsnämnden* dnr 2010/1659) and the radiation protection organization at the Karolinska University Hospital approved this research. Oral and written consents were obtained from the participants. Three families with undiagnosed autosomal dominant ataxia were investigated. Patients, and healthy controls (HC) were evaluated. Age of onset (AO), disease duration (DD) and age of death (AD) were determined. Four affected and four age-matched HC went through structural and functional neuroimaging.

Assessment was performed with Scale for the Assessment and Rating of Ataxia (SARA), Inventory of non-ataxia Symptoms (INAS), Montreal Cognitive Assessment (MoCA), Hospital Anxiety and Depression Scale (HAD-A and HAD-D). In addition, the questionnaire Composite Autonomic Symptom Score (COMPASS 31) was used to screen for dysautonomia (9). Two peripheral nerve biopsies and neuropathological assessment on three patients were carried out.

### Genetic investigations

DNA was isolated from lymphocytes according to standard protocols. The probands in each family had negative genetic screening tests for pathological nucleotide expansions associated with SCA1, 2, 3, 6, 7, 8, 12, 17, DRPLA and SCA31. Linkage to chromosome 16q22.1 was confirmed in family 1. short read (SR) WGS was performed in 3 members of family 1 and 2 each and in the index case of family 3 (Figure 1).

**Figure 1.**
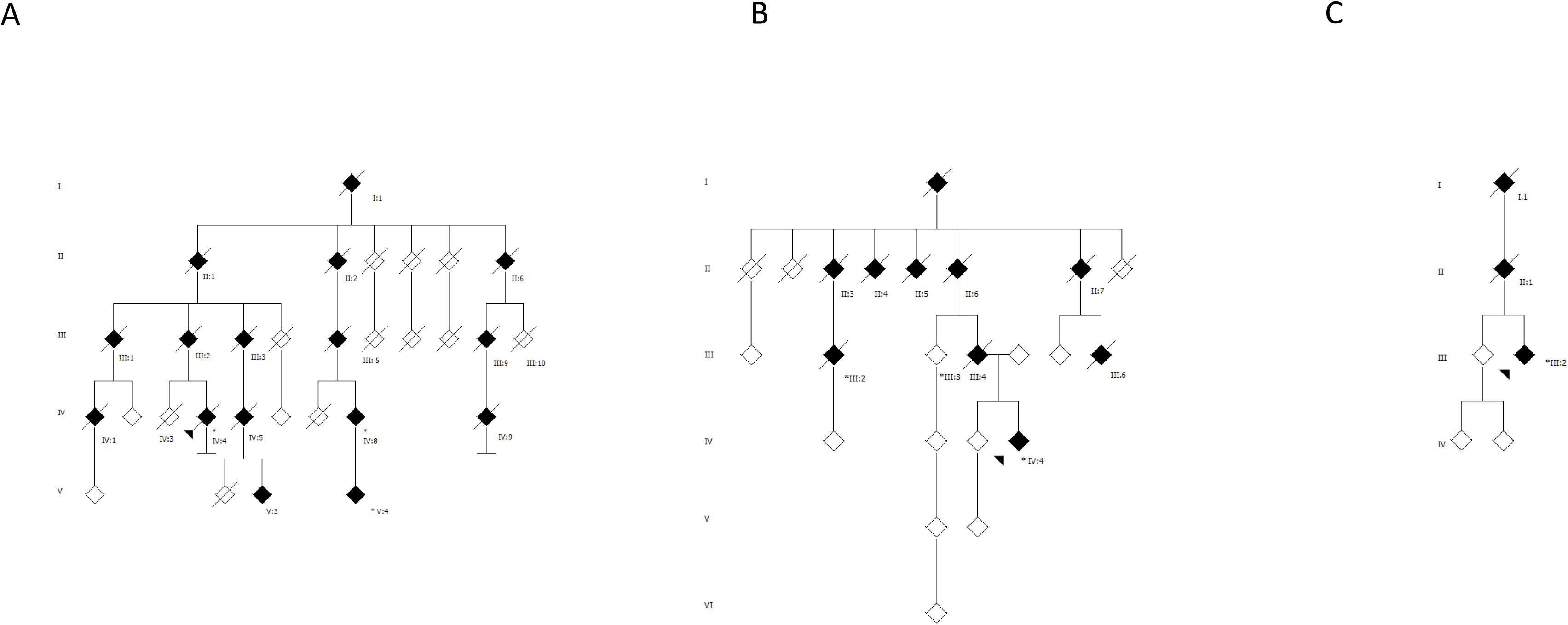
Pedigrees of studied families. Three Swedish families affected by SCA4, anticipation occurs in family 1 and is suggested in the other two families. Mean age of onset was 56.4 years (20-60 years). Family 1 (A) includes 16 patients, of which 7 were examined by the authors. Disease assignment for other 6 patients was corroborated by review of medical notes. Family 2 (B) consists of 10 patients, of which 2 were available for examination. For family 3 (C), 1 patient was examined, disease status in two patients from families 2 and 3 each was assigned based on review of medical records. *These individuals were studied with WGS.

### Repeat expansion detection from WGS data

Expansion analysis was performed using Expansion Hunter *de novo* v0.8.7 in its case-control mode. The hg19 genome data from 1000 SweGen individuals, representing a geographic population cross-section in Sweden was used as a reference cohort (10). Affected family members were used as cases, and unaffected family members were designated additional controls.

### Long read WGS

Cell culture from one affected (Index case in family 3) and one unaffected individual (IV:3 in family 1) was sequenced on Oxford Nanopore PromethION and analysed for the *ZFHX3* STR expansion using IGV.

### Neurophysiological studies

Neurophysiological studies, and tilt test (11) according to previous protocol were applied.

### Biochemistry

Markers for neurodegeneration and monoamine metabolites in the CSF from 2 patients were analysed.

### Neuroimaging

#### Brain, spinal cord and peripheral nerve magnetic resonance imaging (MRI) and analysis

The T1-weighted 3D FSPGR BRAVO sequence was used for MRI volumetry on patients from family 1, for details see supplementary document.

#### PET imaging procedure

All PET measurements were conducted using a high-resolution research tomograph (HRRT) (Siemens Molecular Imaging) after a bolus injection of ^11^C-Flumazenil (375±53 MBq of injected activity) or ^18^F-FDG (247±31 MBq of injected activity).

#### Neuropathology

Neuropathological assessment on patients IV:4 (index case) and IV:4 from family 1 and patient III:2 from family 2 was performed. Examined tissue included cerebrum, cerebellum, brainstem, medulla oblongata, spinal cord, peripheral nerve (sural nerve) and muscle (left anterior tibial muscle). A peripheral nerve biopsy (sural nerve) was also taken from patient IV:1 (family 1).

#### Statistics

Mean age of onset, disease duration, age of death and one way ANOVA tests were performed using SPSS package 20. Significance rates were calculated using the two-tailed, unpaired Student’s t-test with a value of p < 0.05 accepted as significant. When the Levene’s test for equality of variance was significant (p < 0.05) we have reported the results for unequal variance. For the PET data we use two-way ANOVAs (area × genotype) followed by Student’s t-test for pairwise comparisons.

## Results

### Demographics and clinical findings

The three families originate from the same area in Southern Sweden. Families 1 and 2 span six-generations whereas Family 3 spans three generations (Figure 1A-C). The disease affects both women and men. The rate of progression is slow. The mean AO (SD) for all families combined is 56.4 (10.6) years and ranges between 20-60 years (Table 1). Anticipation occurs in all the pedigrees. When a subset of parents (mean AO 56 years) and their children (mean AO 34.4 years), a total of 12 patients with AO documented by neurologists, are compared AO is significantly different (p = 0.035). Disease duration (DD) to death (12 cases) was 27.5 (10.4) years.

**Table 1:**
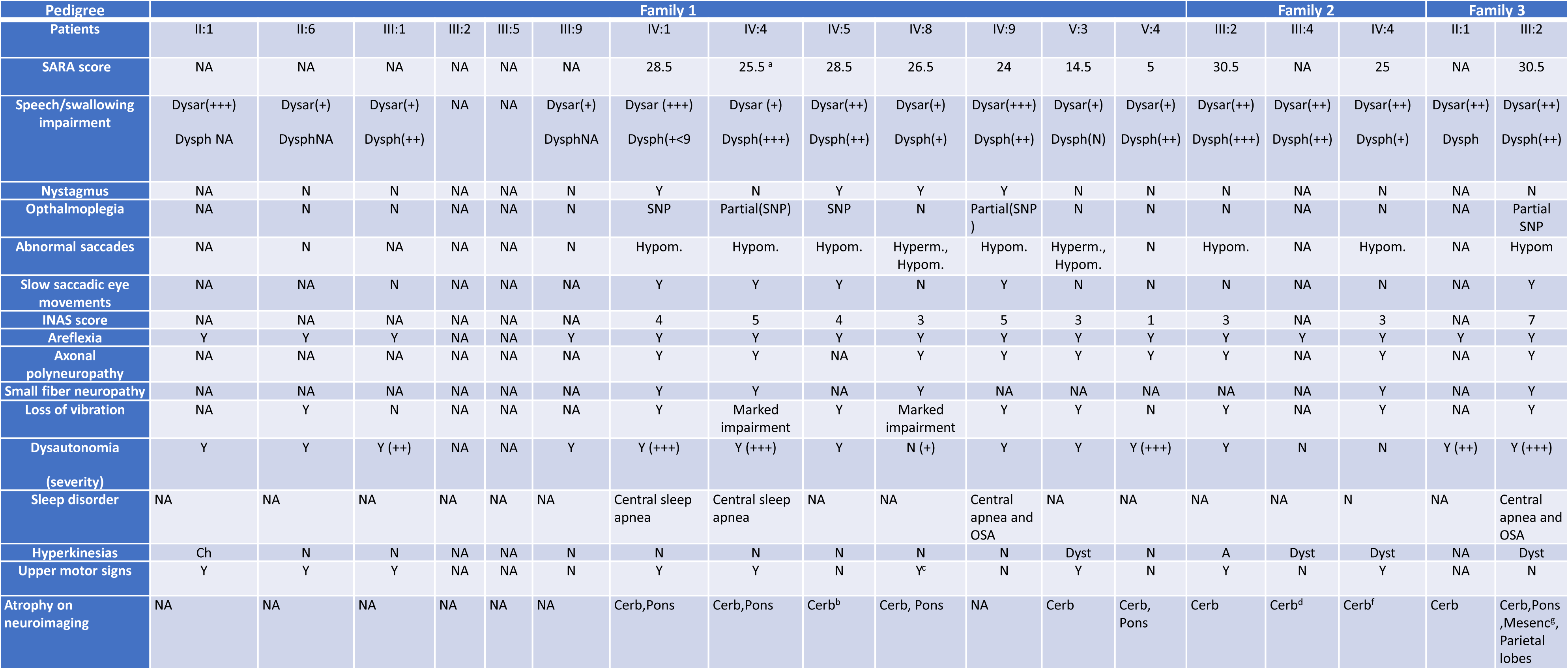
SCA4 pedigree 1 included 16 affected patients, 7 were examined and disease status of other 6 patients was corroborated by review of charts. Disease status for the other 3 was ascribed by history (I:1, II:2 and III:4). ^a^This patient had a unilateral Babinski’s sign before onset of clinical stroke. ^b^Mild atrophy but assessment was carried out with a CT scan. ^c^Babinskis sign during last exam. Pedigree 2 consists of 10 affected patients but only two patients were available for exam. Disease status for the other 7 patients was ascribed by history. ^d^This atrophy was found using a CT scan. ^f^Mild periventricular white matter abnormalities. ^g^Severe atrophy in the mesencephalon. All the examined patients had areflexia and in some cases severe muscle atrophy with contractures. Key: Blepharosp: blepharospasm; Cerb: cerebellum; Female: F; Fasciculat: fasciculations; INAS: Inventory of non-ataxia symptoms; Laryngosp: Laryngospam; M: Male; Minipolym: minipolymyoclonus; Myok: myokymias; NA: not assessed; SNP: supranuclear palsy.

Keeping in with previous descriptions, the phenotype consisted of both cerebellar and sensory ataxia and cerebellar atrophy (4, 6). Several novel features for SCA4 were identified, including dysautonomia, motor neuron affection, eye movement abnormalities (e.g. slow saccades, and ophtalmoplegia) and other movement disorders than ataxia (Tables 1 and 2). In all the examined patients, we found variable dysautonomic features (Supplementary document). Advanced disease was also characterized by severe weight loss, and recurrent pneumonias motivating percutaneous endoscopic gastrostomy (PEG).

**Table 2.** Variable degree of dysautonomia features in three SCA4 families. Key: COMPASS: Composite Autonomic Symptom Score; ESS: Epsworth Sleepiness Scale; N: No; NA: Not assessed. OSA: obstructive sleep apnea; N: No; RR: interval variation on an electrocardiogram; SSR: sympathetic skin response; ^a^ This patient was diagnosed with severe irritable bowel syndrome early in life.

| Pedigree | Pedigree 1 |  |  |  |  |  |  |  |  |  |  |  |  | Pedigree 2 |  |  | Pedigree 3 |
| --- | --- | --- | --- | --- | --- | --- | --- | --- | --- | --- | --- | --- | --- | --- | --- | --- | --- |
| Patients and parameter | II:1 | II:6 | III:1 | III:2 | III:5 | III:9 | IV:1 | IV:4 | IV:5 | IV:8 | IV:9 | V:3 | V:5 | III:2 | III:4 | IV:4 | III:1 |
| Abnormal sweat | NA | NA | NA | NA | NA | NA | Y | Y | Y | N | N | N | N | N | NA | N | N |
| Hot flushes | NA | NA | NA | NA | NA | NA | Y | Y | Y | N | Y | N | N | N | NA | N | N |
| SSR | NA | NA | NA | NA | NA | NA | Absence of reaction | Absence of reaction | NA | Mild abnormality | N | NA | Normal | NA | NA | N | Normal |
| RR variation | NA | NA | NA | NA | NA | NA | Marked reduced | Marked reduced | NA | Reduced | Reduced | NA | Reduced | NA | NA | Reduced | N |
| Orthostatism | Suspected | NA | NA | NA | NA | NA | Y | Y | N | N | Y | N | N | NA | NA | N | Y |
| Pattern of reaction on tilt test | NA | NA | NA | NA | NA | NA | Vasovagal syncope | Asympaticoton | NA | Asympaticoton | NA | NA | Asympaticoton | NA | NA | NA | NA |
| Acrocyanosis | NA | NA | NA | NA | NA | NA | Y | Y | N | N | Y | N | N | N | NA | N | N |
| Urine incontinence (Y/N) | Y | Y | NA | NA | NA | NA | Y | Y | N | N | N | N | N | N | NA | N | Y |
| ESS | NA | NA | NA | NA | NA | NA | 2 | 10 | NA | 4 | 3 | 3 | 0 | 9 | NA | 3 | 12 |
| Sleep registration | NA | NA | NA | NA | NA | NA | Central apnea | Central apnea | NA | N | Central apnea and OSA | NA | N | NA | NA | N | Central apnea and OSA |
| Obstipation | NA | Y | Y | NA | NA | Y | Y | Y | Y<sup>a</sup> | N | Y | N | N | Y | NA | Y | Y |

Areflexia and axonal sensorimotor neuropathy was found in all the affected SCA4 patients examined with ENeG. All five examined patients also had small fiber neuropathy. Notorious motor neuron symptoms and signs occurred across the families and included weakness and the presence of Babinski’s sign. We did not find evidence of seizures, optic atrophy, trigeminal damage, dysmorphism or skeletal malformations. Neither were signs of atrial fibrillation or malignancy. Video recordings displaying phenomenology are provided as supplemental material.

### Neurophysiological studies

RR variability was reduced and the reactions on tilt test were pathological (11), data provided in supplementary material and table 2).

### Biochemistry

Despite widespread neurodegeneration, NfL levels were normal, alpha-fetoprotein (AFP) levels in plasma were also normal in three patients from each family, see supplementary document and table e4.

### Genetic investigations

Multipoint linkage analysis in family 1 confirmed the linkage to chromosome16q22.1 with a maximum LOD score of 3.7 (Figure 2). The linkage peak was located in a 3.69 cM region between markers D16S3086 and D16S512. The whole linked region spanning 25 Mb was custom captured and sequenced in 10 individuals from family 1. No potential SNV or SV pathogenic variant could be identified that segregated with the disease. Based on the sequence data a refined haplotype could be established in the linked region to 23 Mb (flanked by D16S415 and D16S515) (Fig 2A).

**Figure 2.**
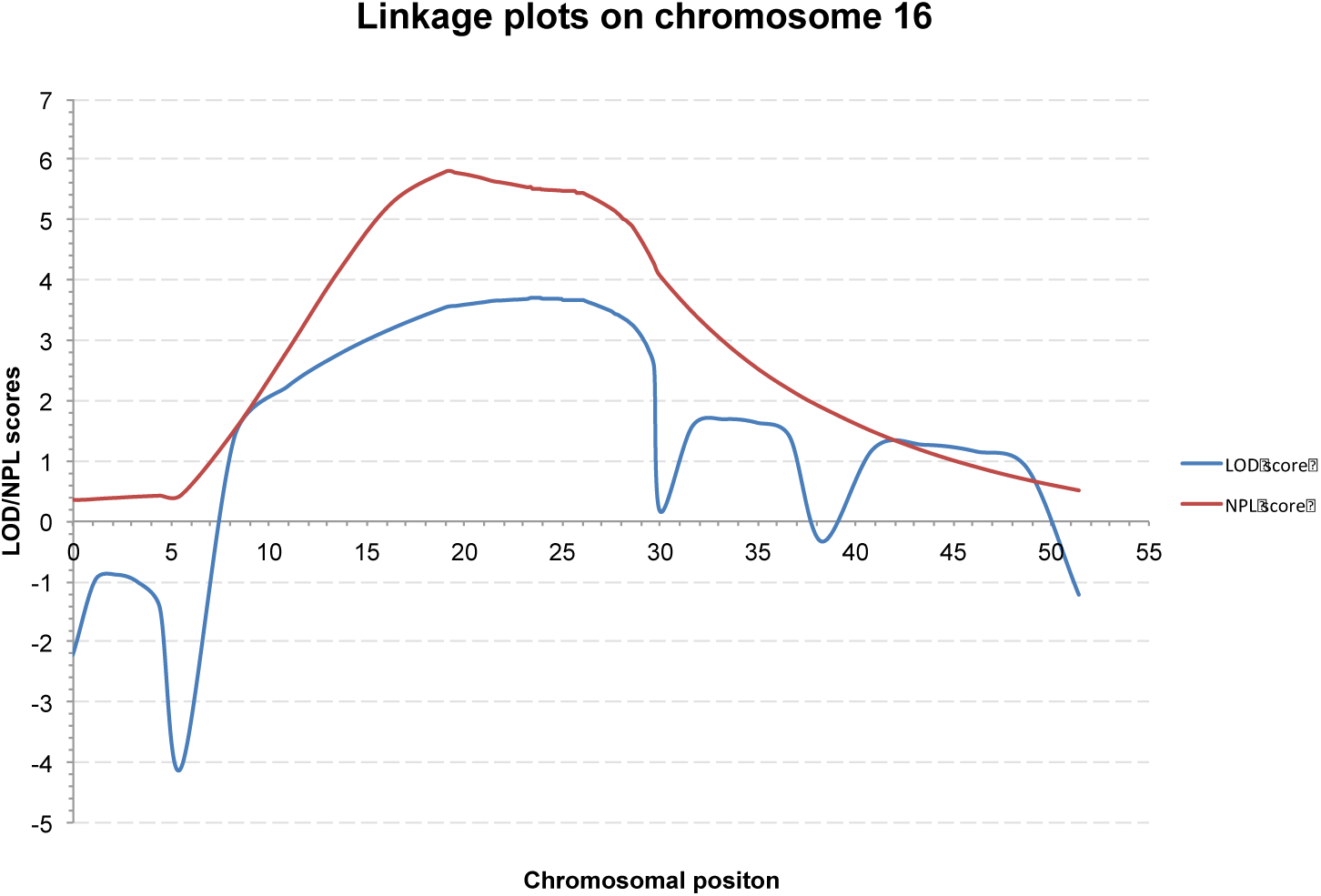
LOD score for SCA4. Multipoint linkage analysis confirmed the linkage with a Maximum LOD score of 3.7 on chromosome 16. Linkage peak was located in a 3.69 cM region between D16S415 and D16S515

### *De novo* repeat expansion analysis

SR WGS was applied to six individuals selected from all three families and subjected to repeat expansion analysis with Expansion Hunter *de novo*. The most significant hit chr16:72821146-72821832 (GGC)n was detected expanded in all five affected individuals, but not in any of the 1001 control samples, including a representative population section plus one unaffected close relative. It was found in the established linkage region. Inspection of alignments in IGV confirm an expanded repeat stretch in the locus (Figure 3A).

**Figure 3.**
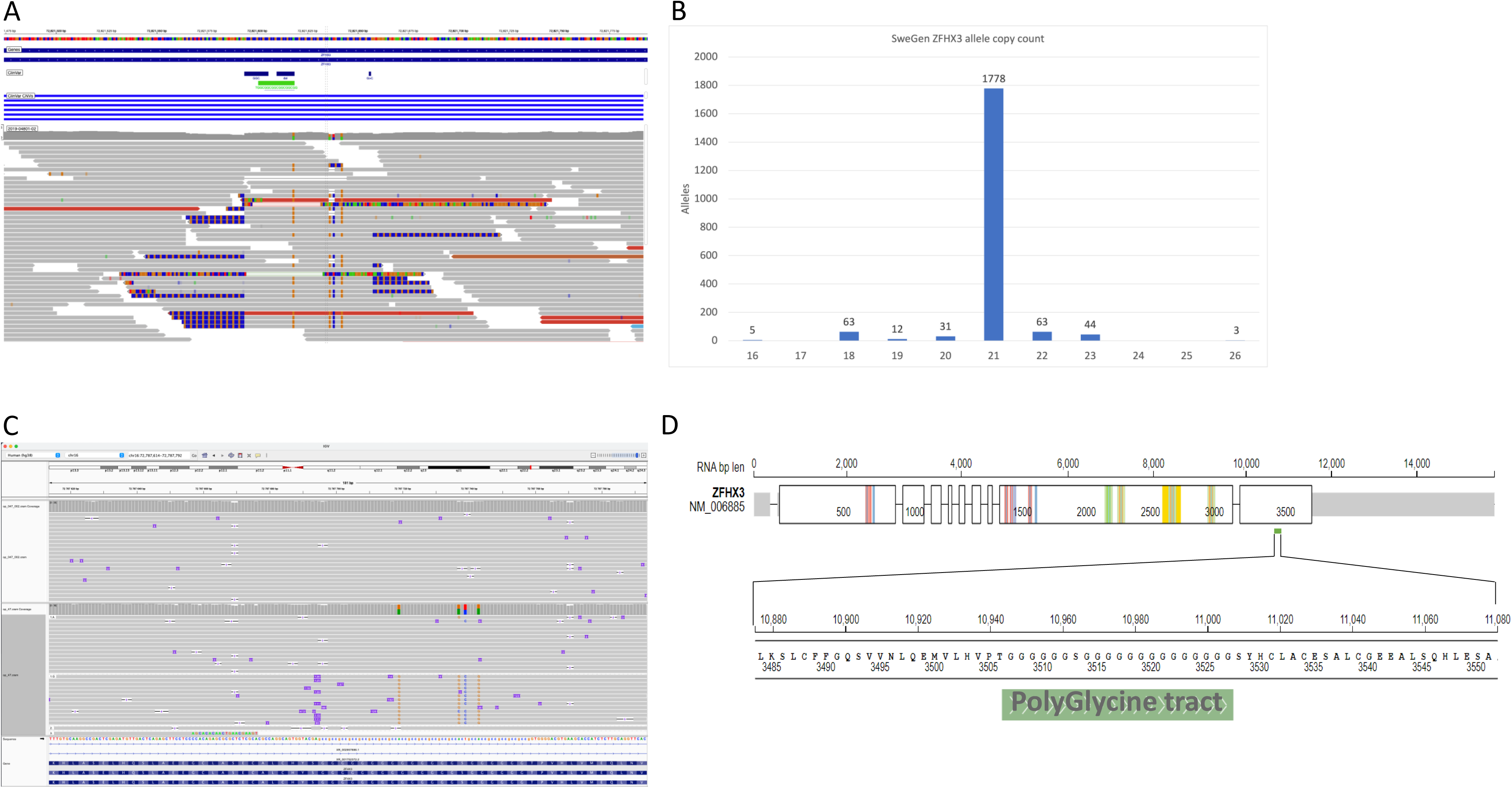
Genome analysis showing a short tandem repeat expansion in the *ZFHX3* gene. A) A) Illumina Short Read Whole Genome Sequencing data from one patient (Family 1, V:4) as visualized in igv.js (Robinson et al 2023). On the top, the reference genome track with the GCC-oligomer reference (GGC in the *ZFHX3* coding direction). Note the soft-clipped expansion sequence bases in blue and orange, with the same repeat unit expanded. Note also the SNV changes in the affected individual compared to the reference repeat, giving a more homogenous repeat without interruptions in the affected individual. A few discordant pairs can be seen, but not as many as would be expected for a considerably larger than insert length expansion, tentatively limiting the total repeat size in affected individuals to below some 80-90 copies. ExpansionHunter genotyping calls this one at 50 copies, but estimation outside read length is known to be approximate. B) Genotyping with ExpansionHunter is considerably more accurate at below read length. Here, 1000 individuals from SweGen (1999 loci, the remaining one typed as 14 copies), with the y-axis describing the number of alleles and the x-axis the number of repeat copies. Expansions are not common in the Swedish population. C) Verification sequencing data with Oxford Nanopore Technologies PromethION displayed with IGV (Robinson 2011). The top track displays an unaffected family member (2021-01634) conspicuously normal in its sequence over the repeat, and the bottom read track an affected individual (2021-24507) with insertions. The reads are grouped according to one of the SNVs changed in the affected individuals, and so shows one normal allele (16 reads indicating on average reference size + 0.5 bp, median 0 sd 3) and one expanded allele (13 reads indicating reference + 120 bp, median 118, sd 21). D) *ZFHX3* gene structure, approximately to genomic scale, with exons shown colored blue, with deep boxes for coding and shallow for non-coding exons, a central line indicating intronic and intragenic sequence. The coding (GGC)n repeat found expanded in affected individuals, but not controls, is highlighted in orange.

### Repeat expansion analysis

ExpansionHunter v4.1 (12) was used to re-detect and preliminarily size expansions in the *ZFHX3* repeat locus (Fig 2B), here defined as hg19 16:72821594-72821656 (GCC)* in the affected families and the SweGen control cohort. The affected individuals were found heterozygous for expansions, indicated as 46-64 copies (Table e3). In other short tandem repeats (STRs), ExpansionHunter will estimate sizes accurately for expansions shorter than read length and give a higher than reference, but not necessarily accurate, repeat copy number count for expansions larger than the read length. The majority of the SweGen loci were detected as 21 copies (1778/1999 called) (average 21, sd 0.8) and the maximum found in the controls was 26 copies. (Figure 3B). The expansion findings in the affected families were otherwise without note, and far from established pathogenicity or perfect segregation.

### Repeat expansion verification

Long read sequencing (LRS) on the index individual from family 3 showed heterozygous expansion at the *ZFHX3* expansion site (Figure 3C): one normal allele (16 reads indicating on average reference size + 0.5 bp, median 0 sd 3) and one expanded allele (13 reads indicating reference + 120 bp, median 118, sd 21).

### Brain and spinal cord MRI

All four patients from family1 who underwent MRI of the brain and spinal cord displayed significant atrophy of the cerebellum, pons, medulla oblongata, and spinal cord (Figure 4). Atrophy of the cerebellum and brainstem was progressive at least in two patients (III:2 and III:4) from family 2.

**Figure 4.**
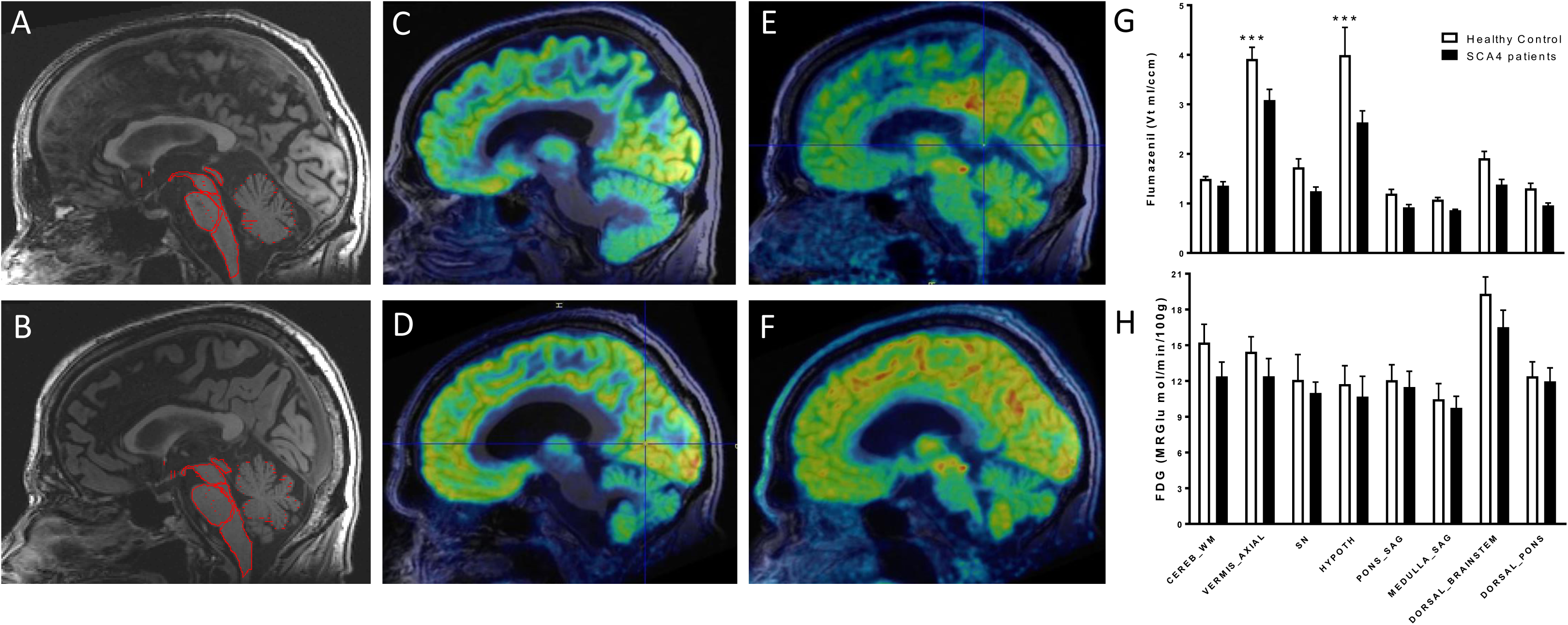
Neuroimaging data in SCA4 patients from pedigree 1. Representative images from patient IV:1 from pedigree 1 (**A, C, E**) and a healthy control (**B, D, F**). (**A, B**) Sagittal T1-weighted brain images in displaying atrophy in the cerebellar vermis, medulla oblongata and pons in patient IV:1. (**C, D)** [C_11_ Flumazenil] V_T_ parametric images of GABA _A_ receptors, to the left subject IV:1 (C) and to the right an age-matched control (D). (**E, F**) FDG SUV/SUVWhite Matter (SUV_ratio) images in patient IV:3 from pedigree 1 (**E**) and a control (**F**). (**G, H**) Degree of Flumazenil and FDG binding by manual ROI in four patients from the SCA4 pedigree 1 and four age-matched controls. Abbreviations: Sup: Superior; Inf: Inferior; A: Anterior; M: Medial; L: Lateral; P: Posterior.

### Peripheral nerve diffusion tensor MRI and nerve magnetisation transfer ratio

Data provided as supplementary material.

### Brain PET studies

Four patients and four sex-and aged matched controls completed both PET studies. Cerebellar PCs express a homogeneous population of GABA_A_ receptors, consisting of α_1_β_2/3_γ_2_ subunits, which bind benzodiazepines (13). Binding of the benzodiazepine ligand, 11C-Flumazenil, was reduced not only in the cerebellum but also in the insula, thalamus and in several brain lobes (frontal, parietal, temporal, limbic and occipital). By manual ROI assessment, this reduction was found in the vermis and hypothalamus. Metabolism measured by 18F-FDG was reduced in SCA4 patients only when the brain regions were pooled together, there was a clear difference in most ROIs but it did not reach statistical significance (Figure 4).

### Neuropathology

The most prominent findings for all three patients were the widespread presence of intranuclear inclusions positive for p62 and ubiquitin; loss of Purkinje cells; loss of motor neurons in the anterior horn of the spinal cord and degeneration of the posterior tracts. The intranuclear inclusions, which appeared eosinophilic in hematoxylin-eosin staining, were round to oval, sometimes ring-shaped, and sometimes there could be up to five of different size in one nucleus. They were found in the brainstem, especially in neurons of the inferior olive nucleus, and in the dentate nucleus of the cerebellum (Figure 5). A few intranuclear inclusions were also detected in the granular cell layer of the cerebellar cortex. Some were seen in motor neurons of the spinal cord. For patient 1 more extensive stainings were performed including sections from the cerebrum. Infrequent neuron in the cerebral cortex also had an intranuclear inclusion. Several inclusions were found in the region of the third ventricle. Inclusions in the epithelium of the choroid plexus, where some had a lentiform shape, were detected as were uncommon inclusions in endothelial cells. Occasional polyG positive intranuclear inclusions were found in neurons of the inferior olive in this patient. From patients 1 and 2 a sural nerve was also examined. Moderate to severe loss of myelinated fibers, both thin and thick, was seen. In the skeletal muscle groups of atrophic fibers were found (Supplementary document).

**Figure 5.**
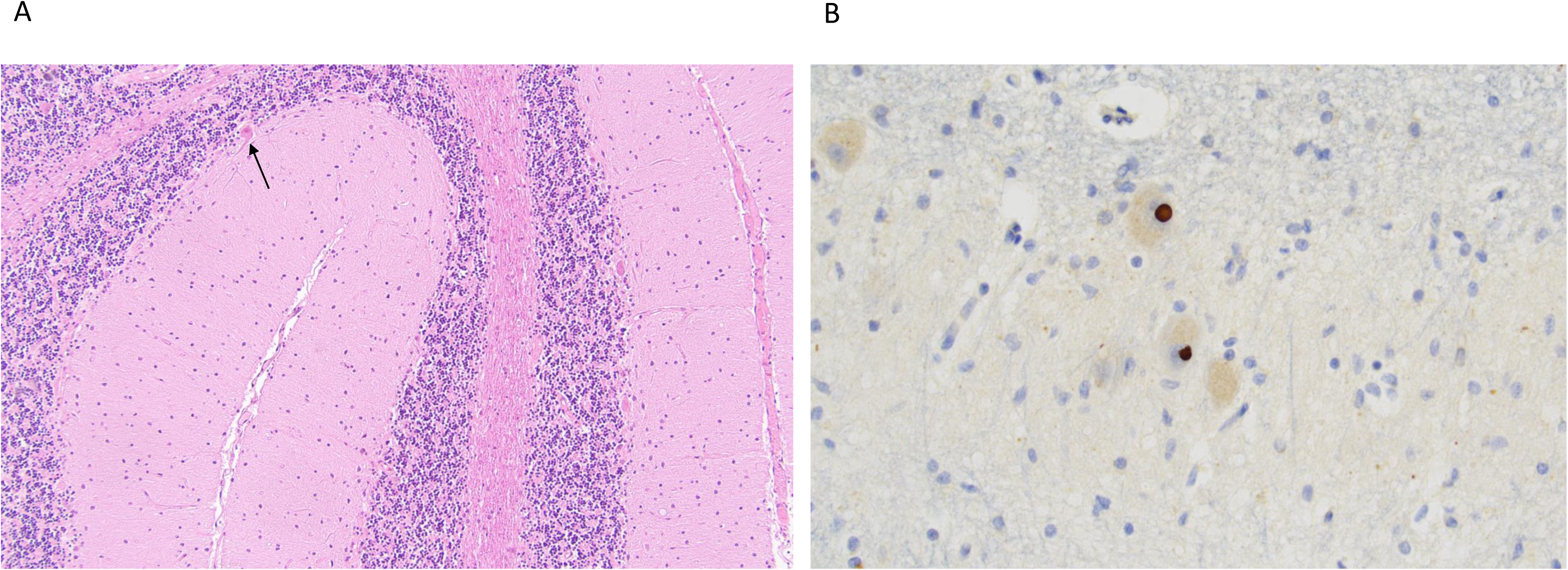
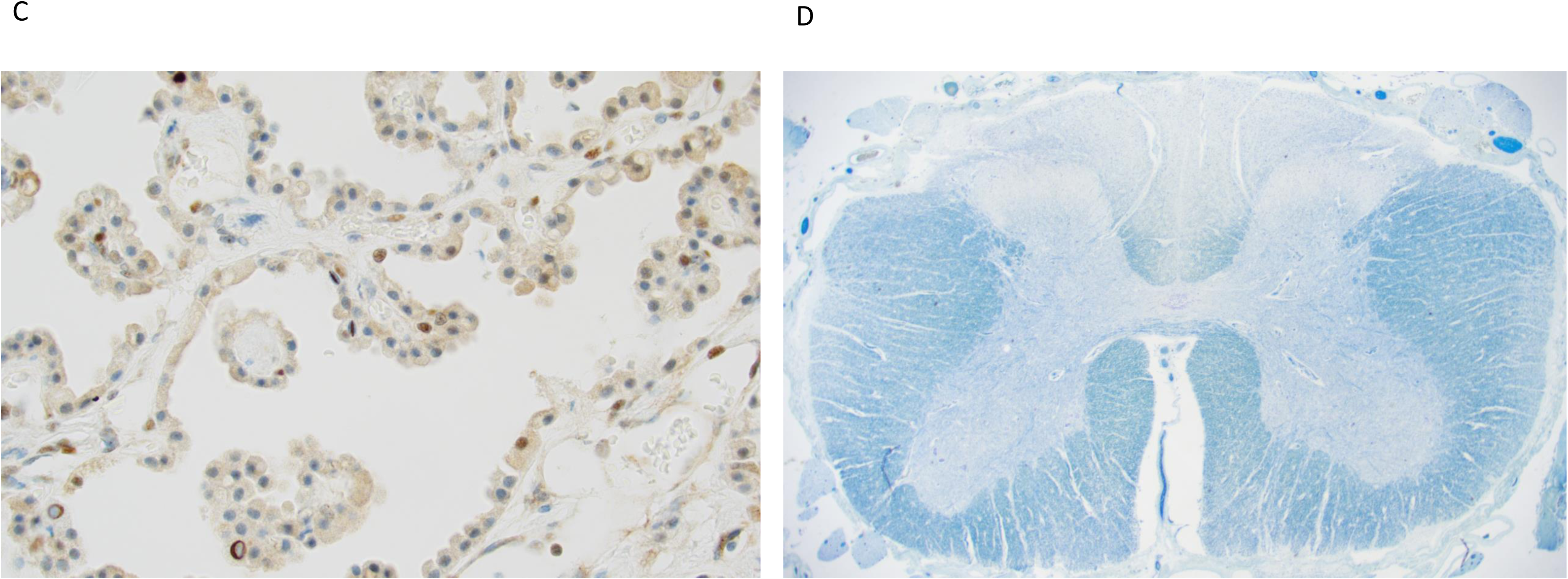
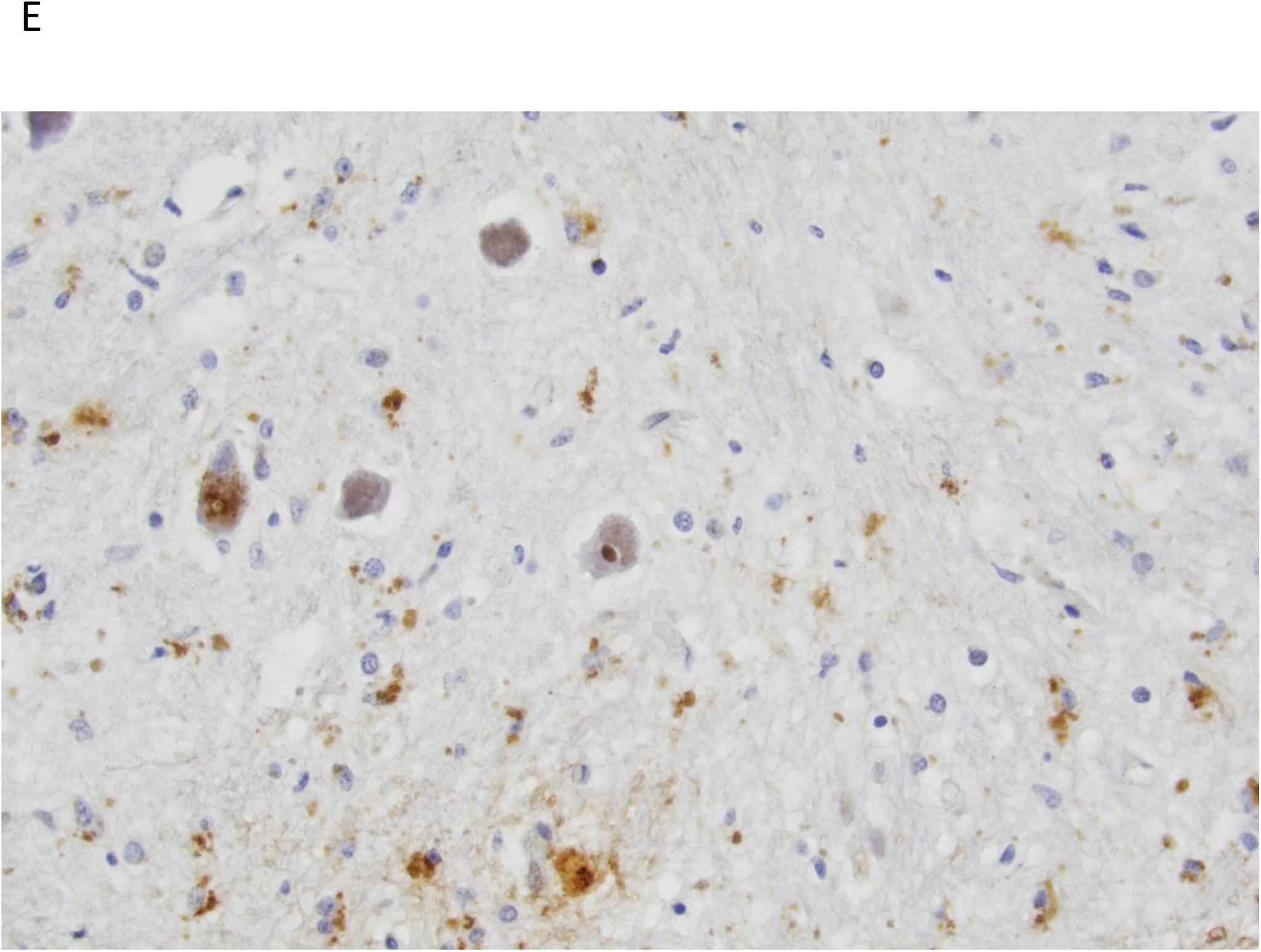
Neuropathology of SCA4. (A) Cerebellum with loss of Purkinje cells. Htx-eosin. (B) Medulla oblongata with the inferior olive nucleus. Intranuclear p62-positive inclusions. (C) Epithelium of the choroid plexus with lentiform intranuclear inclusions (arrow) and an intracytoplasmic inclusion (star). p62. (D) Transverse section of the spinal cord showing loss of motor neurons in the anterior horns and loss of myelinated fibers in the posterior tracts. Luxol fast blue. (E) Medulla oblongata with a polyG-positive intranuclear inclusion in the inferior olive nucleus.

## Discussion

### Disease locus discovery through WGS and STR analysis

Recently, the reduced cost of genome sequencing has triggered a revolution in diagnostics, offering a high diagnostic yield through concurrent testing of multiple rare disease loci in one single assay (14). The availability of innovative callers to determine the presence of expansion in STR loci is of particular interest in ataxia, neurodegenerative disorders and intellectual disability (8), as recent discovery of etiological genes for rare disorders shows (15, 16, 17).

### Extended phenotype of SCA4

These three families expand the SCA4 phenotype, including symptoms beyond ataxia and neuropathy. First, the most striking novel feature is dysautonomia, followed by motor neuron signs, eye movement abnormalities associated with brainstem impairment, dysphagia and dystonia. Second, on MRI imaging we found atrophy in the cerebellum but also in pons, medulla oblongata and spinal cord. Imaging of the peripheral nerves detected reduced maximum FA. In addition, PET imaging revealed reduced 11C-Flumazenil binding in several brain lobes, thalamus, hypothalamus and in the vermis. This reduction may mirror a downregulation of GABA receptors. Third, in neuropathological exams, we found evidence of widespread neurodegeneration in three cases from two different families.

SCA4 is a slowly progressive disease leading to wheel-chair dependency and prostration. Despite widespread neurodegeneration, life expectancy is not reduced and in contrast to other SCAs, CSF-NfL levels were normal (18,19). AO in our families (46.5) is later than in the two previous families (38.3 and 39.3 years, respectively) whereas disease duration is similar to previous descriptions (4, 6).

As in two previously reported SCA4 families, anticipation occurs also in the Swedish families. However, a small sample size and missing data clouds firm interpretations. Our data suggest that earlier AO may be associated with more severe dysautonomia. Expansion in the *ZHFX3* locus is consistent with anticipation and would have been more difficult to explain from an SNV finding. We suspect that as more samples undergo long read sequencing, a gradual size increase of repeat expansion with generation and disease severity will be seen. Scandinavian heritage in the first reported SCA4 family raises the question of a founder effect, when taken together with the families reported here, although expansion is rare in the population (not in 1000 Swedish controls; Figure 3B).

Polyneuropathy is a common feature in SCAs, but its frequency varies depending on the SCA subtype ranging from 63-82% in the polyQ SCAs (20). Our results indicate that polyneuropathy is a universal feature in SCA4. The fact that areflexia was noticed in the proband of family1 in his 20s suggests that subclinical neuropathy was already present. This trait has been proposed as the earliest sign of disease in the North American family (4). We also found small fiber neuropathy but no signs of trigeminal dysfunction as reported previously (6). Small fiber neuropathy is a well characterized cause of autonomic dysfunction (21). However, the fact that four patients suffered from central sleep apnea argues against this abnormality as the only contributing factor to dysautonomia. Reduced 11C-Flumazenil binding may be explained by either reduced expression of GABA receptors or loss of GABAergic neurons in the hypothalamus. Autonomic dysfunction is not a unique feature of SCA4, but in no other SCA is this feature so predominant or the presenting symptom. Dysautonomia is also common in the polyQ SCAs (22, 23, 24).

Only one earlier neuropathological description in a SCA4 case exists limited to the study of the cerebellum and brainstem only (7). Besides demyelinization of cerebellar and brainstem fiber tracts, severe loss of Purkinje cells and neuronal loss in several brainstem nuclei (both cranial nerve nuclei, SN and the inferior olive) was found (7). In addition, no signs of neuronal intranuclear inclusions (NII) were found when IC2 antibodies were used (7). Only one of our cases had neuronal loss in the hypoglossus nuclei. We found evidence of widespread neurodegeneration beyond those regions in three patients from two SCA4 families. Loss of lower motor neurons found in all 3 patients who displayed variable degrees of muscle atrophy, weakness, and/or fasciculations. Several features and observations argue strongly against SCA4 being the same ataxia disease described by A. Biemond (5) (Extended discussion is provided in supplementary material).

### Disease locus discovery

The STR locus implicated in this study was in the coding region of *ZFHX3* (Figure 3D), encoding a glycine repeat, which partly explains why it has escaped earlier attempts. The gene had previously been considered in the etiology of SCA4 on account of also holding one of the nine (CAG)n stretches then investigated (6).

*Zhfx3* is expressed in the developing mouse brain (25). The protein was most intense at embryonic day 20 and highly expressed in the midbrain and diencephalon whereas mRNA was highly expressed in in the brainstem (during embryogenesis and in neonatal brains), the major site of expression was in postmitotic brainstem neurons (26). Outside the CNS, IHC staining was found in the nuclei of immature neurons (adrenal medulla and nerve ganglion) and in pulmonary smooth muscles (26). *Zhfx3* is highly expressed in the suprachiasmatic nucleus of the hypothalamus (27), induced missense variant in *zfhx3* accelerates circadian rhythms in mice (28).

Expanded protein coding polyglycine stretches have not previously been described in disease, although the *C9orf72* ALS locus shows a poly-glycine-alanine-expansion in affected individuals (29). The presence of nuclear inclusions in the patient are reminiscent of neuronal intranuclear inclusion disorder disease (NIID), associated with a GGC repeat expansion in the 5’ untranslated region of the *NOTCH2NLC* gene (30, 31, 32). Translation of upstream open reading frame GGC expansions has been seen in NIID and found to accumulate into inclusions in mice and cell models (33).

The *zfhx3* gene has been studied in zebrafish as a candidate in the Undiagnosed Diseases Project, where its homolog was found to be strongly expressed during neurogenesis in regions with roles in motor control and coordination and point mutations were found in rare disorder cases with epilepsy (34). Biallelic *zfhx3* knockout is lethal in mice, but heterozygous knockouts display an increased pre-waining mortality (35). Similarly, mice with a heterozygous N-ethyl-N-nitrosourea (ENU) induced null mutation barely survive to weaning, whereas its homozygous is also lethal (28). In addition, *ZFHX3* plays at least three different known roles in malignancy (36, 37, 38), acting as a tumor suppressor in the prostate (39). Variants in *ZFHX3* are associated with increased risk for Kawasaki disease, atrial fibrillation and with coronary heart disease among Asians (40, 41, 42, 43, 44, 45). None of the patients we report here had malignancy, seizures or atrial fibrillation.

Of note are the commonly occurring serine interspersed in the poly-glycine stretch, and so AGT codon interrupting the (GGC)n, and occasionally GGT, also coding for glycine. In the expanded variants the repeat is more homogenous, with fewer interruptions that could assist polymerase fidelity and inhibit STR prolongation. SCA4 indeed shows signs of anticipation. With 99,8% of control loci below 24 copies, loci with 25-42 copies may represent pre-mutations.

In conclusion, SCA4 is a neurodegenerative disease underlined by intranuclear neuronal inclusions and a novel GCC expansion in *ZFHX3*. By means of WGS we were able to identify an underlying mutation perfectly segregating with autosomal dominant inheritance in three kindreds, and consistent with anticipation. We present the first coding region poly-glycine expansion disorder and show how WGS in well studied families can support disease locus discovery also in a long-established clinical entity with previously unknown genetic etiology.

## Data Availability

All data produced in the present work are contained in the manuscript

## Abbreviation

SCA4: Spinocerebellar ataxia type 4

## Acknowledgments

We are in deep debt with all the patients and their relatives for their kind participation. We are also grateful to Dr T. Karu for her kind assistance with the assessments at the physiology department in our hospital. We would like to thank UPPMAX for the use of computer infrastructure and the support from the National Genomics Infrastructure Stockholm, Uppsala Genome Center and Clinical Genomics Stockholm facility at the Science for Life Laboratory in providing assistance in massive parallel sequencing. The study was supported by the Stockholm City Council. AW and PS are Wallenberg Clinical Scholars. MP’s research is supported by Region Stockholm.

## Notes

### Competing Interest Statement

The authors have declared no competing interest.

### Author Declarations

The Swedish Ethical Review Authority

