## Supplementary document for "Spinocerebellar ataxia type 4 is caused by a GGC expansion in the *ZFHX3* gene and is associated with prominent dysautonomia and motor neuron signs"

### Samples and methods

### 1. Clinical data

Evaluations were pursued as described in the main text and summarized in Tables 1 and 2. Our findings were compared to phenotypes were compared to two previous publications on spinocerebellar ataxia 4 (SCA4) (1, 2).

### 2. Neurophysiological studies

Electroneurography (ENeG), electromyography (EMG), quantitative sensory test (QST), variability of RR-interval, skin sudomotor response (SSR), ambulatory polysomnography with Embletta equipment, hearing test were applied according to standard protocols. Tilt test was performed and evaluated according to Sutton et al (3).

### 3. Neuroimaging

#### 1.1 The spinal cord and peripheral nerve MRI:

Patients IV:1, IV:4, IV:5 and V:4 from family 1 were studied with MRI of the spinal cord and peripheral nerves. Those structures were examined on a 3.0 Tesla MR scanner (Magnetom VERIO, Siemens Healthcare) with a 32-channel spine coil and a transmit/receive 15-channel knee coil. The spinal cord was examined with a sagittal T2-weighted sequence [TR=3000 ms, TE=102 ms, FOV=350 mm, acquisition matrix=512×384, slices=15, slice thickness/gap=3.0/0.3 mm, voxel size=1.0×0.8×3.0 mm, NEX=2]. The left distal thigh was examined with a 3D T2-weighted sequence [TR=1500 ms, TE=134 ms, FOV=178 mm, acquisition matrix=192×188, slices=96, slice thickness=1.0 mm, voxel size=0.9×0.9×1.0 mm, NEX=2], a single-shot diffusion-weighted EPI sequence with a b value of 1000 s/mm$^2$ in 30 diffusion encoding directions plus a reference image without diffusion-weighting [TR=3000 ms, TE=85 ms, FOV=155 mm, acquisition matrix=124×124, slices=10, slice thickness/gap=5/5 mm, voxel size=1.2×1.2×5.0 mm, NEX=6, spectral adiabatic inversion recovery (SPAIR) was used for fat suppression plus saturations band (regional saturation technique; REST) anterior and posterior over the subcutaneous tissue, partial Fourier encoding=5/8, parallel imaging (GRAPPA) with acceleration factor=3] with calculation of fractional anisotropy (FA) maps on a voxel-by-voxel basis, and finally with magnetisation transfer (MT) imaging without (MTnonsat) and with an off-resonance saturation pulse (MTsat=1.5 kHz off-resonance, 10 ms long Gaussian pulse, 5000 effective flip angle) [TR=33 ms, TE=7 ms, FOV=154 mm, acquisition matrix=192×192, slices=20, slice thickness/gap=5.0/1.0 mm, voxel size=0.8×0.8×5.0 mm, flip angle=10, NEX=5].

**Data analysis brain MRI**

The T1-weighted 3D FSPGR BRAVO sequence was used for MRI volumetry. Using software tools from the FMRIB software library (http://www.fmrib.ox.ac.uk/fsl), the brain was extracted from surrounding bone, and segmented into brain tissue and cerebrospinal fluid. The brain was manually segmented into cerebrum, mesencephalon, pons, medulla oblongata and cerebellum (Schulz et al., 2010). The percentage of each structure's volume, as part of the total brain volume, was calculated. The midsagittal distance of the spinal cord, at the midlevel of each vertebra from C1 to Th12, was measured and a mean value was calculated for each patient. Using T2-weighted images for anatomical reference, a circular region of interest (ROI), approximately 6 mm in diameter, was placed in the sciatic nerve on fractal anisotropy (FA) maps. The highest value was recorded on each slice and a mean value was calculated for each patient. On magnetisation transfer (MT) images, without and with a saturation pulse, a ROI was placed in identical positions in the sciatic nerve. Due to proximal and distal fold-over artifacts in the MT images, the central 13 slices were used for analysis. Magnetisation transfer ratio (MTR) was calculated MTR=100*(MTnonsat-MTsat)/MTnonsat, and mean MTR was calculated for each patient. Finally, a circular ROI, approximately 10 mm in diameter, was placed in the vastus medial muscle and mean MTR was calculated for each patient.

**PET imaging procedure**

All PET measurements were conducted using a high-resolution research tomograph (HRRT) (Siemens Molecular Imaging) after a bolus injection of $^{11}$C-Flumazenil (375±53 MBq of injected activity) or $^{18}$F-FDG (247±31 MBq of injected activity). A 6-minute transmission scan using a $^{137}$Cs source was first acquired for attenuation correction. Emission data for both radioligands were acquired in list mode for a period of 65 minutes. Dynamic images were reconstructed in a series 32 time frames using three-dimensional ordinary Poisson ordered subset expectation maximization (OP-3D-OSEM) including modeling of the system´s point spread function (PSF) with an estimated resolution around 2 mm (4). To prevent head motion during the PET measurement, an individual plaster helmet was made for each patient and healthy control (5). Images were also corrected for motion with a post-reconstruction frame-to-frame correction realignment algorithm as previously described (6). $^{11}$C-Flumazenil PET studies were analyzed with Logan invasive and with arterial blood input function. The outcome measure was Total Distribution Volume ($V_T$). $^{18}$F-FDG PET studies were analyzed with the Patlak plot. The outcome measure was the regional metabolic rate of glucose (MRGlu). All kinetic modeling was performed using PMOD v3.3 (PMOD Group, Switzerland). Individual voxel-based $V_T$ images were generated using the WAPI software (7) with optimized parameters for HRRT as previously reported (6). 3D T1-weighted MRI images were first realigned along the plane connecting the anterior and the posterior commissures and then coregistered to the summed PET images using SPM5 (Statistical parametric mapping, Wellcome Trust Centre for Neuroimaging, U.K.). ROIs were manually delineated on patients´s MRI on sagittal midbrain, dorsal brainstem, dorsal pons, sagittal colliculi, sagittal midbrain, sagittal medulla, axial cerebellar vermis, cerebellar white matter, red nucleus and using the software Human brain atlas (Hba). The AAL (automatic anatomical labeling) template was used to define other encephalic regions of interest: cerebellar grey matters, thalamus, caudate, putamen, pallidum, insula, amygdala, hypothalamus and frontal cerebral cortex (8). ROIs were then applied to the dynamic PET images using the MRI-PET transformation matrix to obtain regional time activity curves (TACs). The generated volume ($mm^3$) of the ROIs was also used to perform a volumetric analysis of all the defined regions in SCA 4 patients and healthy controls.

**Biochemistry**

CSF analyses were performed in 2 patients from family 1. Levels for different biomarkers of neurodegeneration, GFA-p, S-100 and monoamine metabolites in the CSF were determined. Tissue punches from several parts of cerebral cortex along with hippocampus, thalamus, hypothalamus, nucleus caudatus, cerebellum, pons, DRG, peripheral nerve and the spinal cord were microdissected out from fresh-frozen postmortem coronal sections from the index cases of family 1, patient III:2 of family 2 and two matched controls. Frozen tissue was sonicated in 1% SDS and boiled. Protein concentration was determined using the Pierce bicinchoninic acid protein assay method (Thermo Fisher Scientific). Equal amounts of protein were separated by SDS/PAGE and transferred to Immobilon-P (PVDF) membranes. Immunoblotting for GAD65 (Chemicon; dilution 1:1000) was carried out in tissue punches from the brain, spinal and peripheral nerve.

#### *4. Genetic analyses*

##### *4.1 Linkage analysis*

Linkage analysis was focused on chromosome 16, a SCA4 candidate region from previous reports (1,2). Twenty microsatellite markers covering about 50 cM were selected for genotyping in 10 members from the large Swedish SCA4 family (Family 1). Genotype errors were checked prior to linkage analyses by using PedCheck (3). Markers were removed when genotype errors were identified for the corresponding subjects. GENEHUNTER v. 2.1 (4) was used for both parametric and non-parametric linkage analysis. Parametric linkage analysis was performed assuming an autosomal dominant model with a penetrance of 0.999 for heterozygotes, disease-allele frequency of 0.001, and phenocopy rate of 0.001. LOD scores were calculated under dominant model. Both multipoint and single point analysis were performed. The positions of marker loci were based on published human genetic maps (National Center for Biotechnology Information [NCBI]). Linkage to chromosome 16q22.1 was confirmed in family 1 using markers D16S3031, D16S3019, D16S397, D16S3067, D16S3141, D16S496, D16S3085, D16S3107, D16S421, D16S3086, D16S3095, D16S3624, D16S3059, D16S512, D16S3018, D16S516 and D16S402.

##### 4.2 Targeted region short read sequencing

***Target capture kit design and sequencing and whole genome sequencing***

Targeted sequencing focused on a 23 Mb genomic region which covered the entire linkage peak on chromosome 16 (53633818-76593135 GRCh37/hg19,(Figure 2 in main text). Enrichment of the target region was reached by using a custom-designed NimbleGen SeqCap EZ Developer library (Roche NimbleGen, Inc. Madison, WI 53719 USA).

Ten DNA samples, four unaffected and six affected from family 1 (IV:1, IV:4 and healthy sibling, IV:5 and healthy daughter, IV:8, V:4, and IV:9 with healthy parent and relative) were selected for targeted sequencing (custom capture) (Figure 1). The sequencing work was done at SciLifeLab Stockholm, Sweden. Each DNA library was prepared from 100 ng of genomic DNA. DNA was sheared to 300 bp using a Covaris S2 instrument and enriched by using the custom-designed NimbleGen SeqCap kit. Clustering was performed on a cBot cluster generation system and sequencing was performed on HiSeq 2500 with 2x 100 paired-end reads setting (targeted sequencing) according to manufacturer's instructions or HiSeqX (HiSeq Control Software 3.3.76/RTA 2.7.6) with a 2x151 setup using 'HiSeq X SBS' chemistry (whole genome sequencing). Base conversion was done using CASAVA v1.8.2. The quality scale is Sanger / phred33 /Illumina 1.8+. All samples yielded the expected number of sequencing reads.

***Sequencing data analysis***

Sequencing reads were mapped against the human reference genome assembly hg19/GRCh37 by using Burrows-Wheeler Aligner (bwa version 0.7.12, http://bio-bwa.sourceforge.net/, (9). Reads were trimmed by a quality score of 20. Sequence variants were called by using Genome Analysis Toolkit/UnifiedGenotyper (GATK, version 3.0-0, https://www.broadinstitute.org/gatk/), (10). Local realignment and base recalibration were applied before variant calling. PCR duplicates were removed using Sequence Alignment/Map (SAM) Tools version 0.1.19 (http://samtools.sourceforge.net/) prior to variant calling. The resulting variants were annotated using Annovar (http://www.openbioinformatics.org/annovar/) (11), and SNP142 (http://hgdownload.soe.ucsc.edu/goldenPath/hg19/database/); the 1000 genome project (http://www.1000genomes.org/) were used for common variants filtering. Functional roles of variants were annotated based on integration of varieties of reference resources such as refGene (http://hgdownload.soe.ucsc.edu/goldenPath/hg19/database/) and predication tools via Annovar.

**Variant filtering and Sanger sequencing validation**

As a basic quality control step, initially called variants were filtered first based on the read depth. Variants with a depth less than 10 X or larger than 1000 X were excluded in the further analysis process.

Interesting variants were selected by using several filtering criteria: co-segregation with the disease; rare, i.e. with a minor allele frequency (MAF) ≤ 1 % in general populations of the1000 genome project (12); and with a higher functional impact.

**4.3 Short read whole genome sequencing**

Whole genome sequencing was performed in 3 members of family 1 and 2 each and in the index case of family 3 and analysed as previously described (13). Briefly, sequencing libraries were prepared from DNA from whole blood, and subjected to paired end short read sequencing on Illumina instruments to approximately 30x coverage. The resulting data was analysed bioinformatically for single nucleotide, structural and repeat expansion variants and triaged A) for clinically relevant findings in the entire genome and B) variants of interest in the linkage region on chromosome 16q22.1.

**4.4 Repeat expansion detection from short read WGS data**

Expansion analysis was performed using Expansion Hunter *de novo* v0.8.7 in its case-control mode. The hg19 genome data from 1000 SweGen individuals, representing a geographic population cross-section in Sweden was used as a reference cohort (14). Affected family members were used as cases, and unaffected family members were designated additional controls. Well established genes with known nucleotide repeat expansions were analysed as previously reported using Expansion Hunter (13, 15), with the addition of a tentative expansion definition of the *ZFHX3* locus. The same method was applied to the SweGen cohort.

**4.4 Long read whole genome sequencing**

Cell culture from one affected individual and one unaffected individual (Table e3) was grown in a 75 cm2 culture bottle, harvested and snap frozen in liquid nitrogen, DNA prepared according to manufactures instruction using Monarch cell extraction kit, resulting about 3.3 ug DNA, short fragments eliminated with Circulomics SRE kit and sequenced on Oxford Nanopore PromethION by the Science for Life Laboratory National Genome Infrastructure Uppsala Genome Center. 7.94 M and 10.1 M reads were generated with R9 flowcell chemistry for a total of 119.2 Gb and 88.25 Gb passed bases with high-accuracy base calling and failing reads below an average Phred score quality of 10. Read length N50 was estimated at 24.6 kb and 13.5 kb after outliers were discarded. IGV was used to visualise the ZFHX3 expansion locus.

#### 5. Neuropathology

After fixation in formaldehyde material for histological examination was collected according to a protocol for neurodegenerative diseases. For patient 1 tissue from the region around the third ventricle was also sampled. For all patients the spinal cord was included. Stainings on 5µm thick sections were made with haematoxylin-eosin, Luxol fast blue and Bielschowsky silver stain. Immunostainings were performed on numerous sections with antibodies against ubiquitin and p62. Furthermore, staining with antibody against polyG was performed on sections from patients IV:4 from family 1.

### Results

#### 1. Clinical data

Anticipation was documented in the Swedish SCA4 families. One patient affected by ataxia also had a history of chronic alcohol abuse and was not included in the analysis. None of the other ataxia patients had ever abused alcohol or been treated with antiepileptic drugs or cytostatic agents. In all the examined patients, we found variable dysautonomic features which included neurogenic orthostatism, obstipation, abnormal sweating, hot flushes, sleep abnormalities, acrocyanosis, urinary incontinence and/or erectile dysfunction. Recurrent syncope due to severe orthostatism was the initial symptom in the index case in family 1, it also affected another patient in family 2. Early erectile dysfunction was also common, in one case present since adolescence and prodromal to motor onset. One patient/ in family 1 had heart failure with no records of sleep apnea. Central sleep apnea was found in 4 patients without evidence of underlying heart disease (Table 2). In contrast with previous reports, dysarthria and dysphagia were common (16/18) even though severity was variable. Advanced disease was also characterized by severe weight loss, and recurrent pneumonias motivating percutaneous endoscopic gastrostomy (PEG).

Areflexia and axonal sensorimotor neuropathy was found in all the affected SCA4 patients examined with ENeG. All five examined patients studied with QST also had small fiber neuropathy. Neuropathy was severe and disabling in one case as it overshadowed ataxia (IV:9). In another patient areflexia was noticed in his 20s, many years before onset of ataxia. All the examined patients complained of worsened balance when exposed to darkness. Loss of vibration was also very common, agraphestesia occurred in those with advance neuropathy. Notorious motor neuron symptoms and signs occurred across the families and included weakness (advanced disease), fibrillations, myokymias and the presence of Babinski's sign (Table 2). Besides cerebellar eye movement abnormalities, abnormalities associated with brainstem dysfunction such as slow saccades and variable ophthalmoplegia were found in those with advanced disease. We also documented dystonia and mirror movements. Two patients in family 2 presented with dystonia; one of them (IV:4) had dystonia in the diaphragm, eyelids, extremities and laryngospasm. Her father did suffer from dystonia too (III:4). Dystonia occurred to a much lesser degree only in one patient in family 1, chorea was reported in medical notes only one patient (II:1 in family 1) but no video recording documenting it was available.

Dysautonomia features in these families includes neurogenic orthostatism, obstipation, abnormal sweating, hot flushes, sleep abnormalities, acrocyanosis, urinary incontinence and/or erectile dysfunction. Recurrent syncope due to severe orthostatism was the initial symptom in the index case in family 1, it also affected another patient in family 2. Early erectile dysfunction was also common, in one case present since adolescence and prodromal to motor onset. One patient in family 1 had heart failure with no records of sleep apnea. Central sleep apnea was found in 4 patients without evidence of underlying heart disease (Table 2). In contrast with previous reports, dysarthria and dysphagia were common (16/18) even though severity was variable. We also documented dystonia and mirror movements. Cognitive decline became evident in two patients only, the index case from family 1 after a stroke episode and in one patient from family 2 at advanced age. The latter had neuropathological features compatible with Alzheimer's disease. Another patient in family 1 had delirium secondary to subdural hematomas, he recovered gradually with conservative treatment.

Hearing loss was present in four patients in family1 and in one patient from family 2. However, this deficit, found with advanced age, was compatible with presbyacusis. Only two patients required treatment with hearing aid devices, in one case the hearing deficit was asymmetric. We did not find evidence of seizures, optic atrophy, trigeminal damage, dysmorphism or skeletal malformations. Neither were signs of atrial fibrillation or malignancy.

**Peripheral nerve diffusion tensor MRI and nerve magnetisation transfer ratio**

One patient (IV:4) had its division of the sciatic nerve, into the common peroneal and tibial nerve, proximal on the thigh and therefore the broader tibial nerve was examined. Patient IV:8 was excluded since the distal thigh did not fit into the coil. In the other two participants the sciatic nerve was examined. In all three studied patients, maximum fractional anisotropy (FA) was significantly lower than in controls. Additional neuroimaging data is presented in table e4.

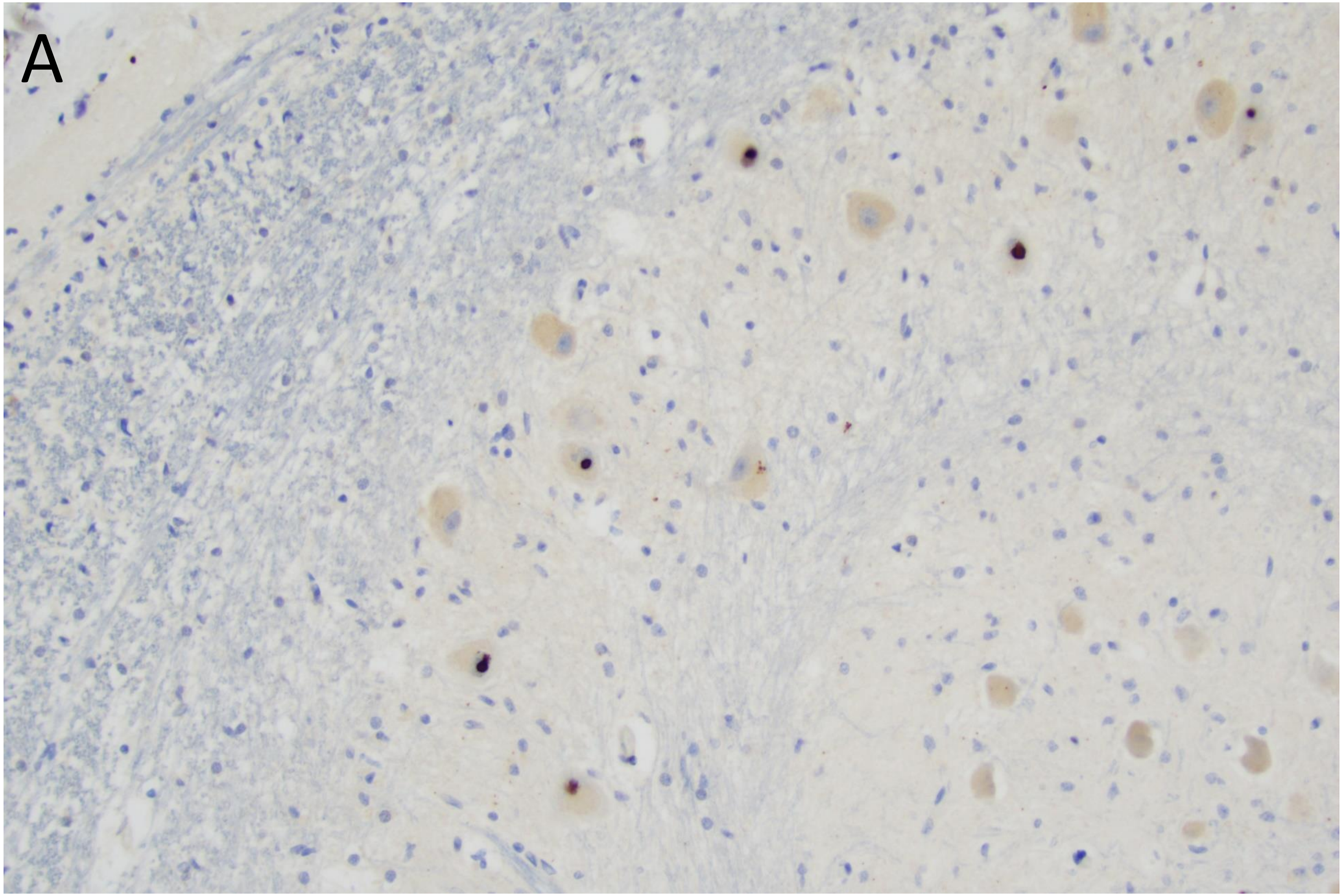
A

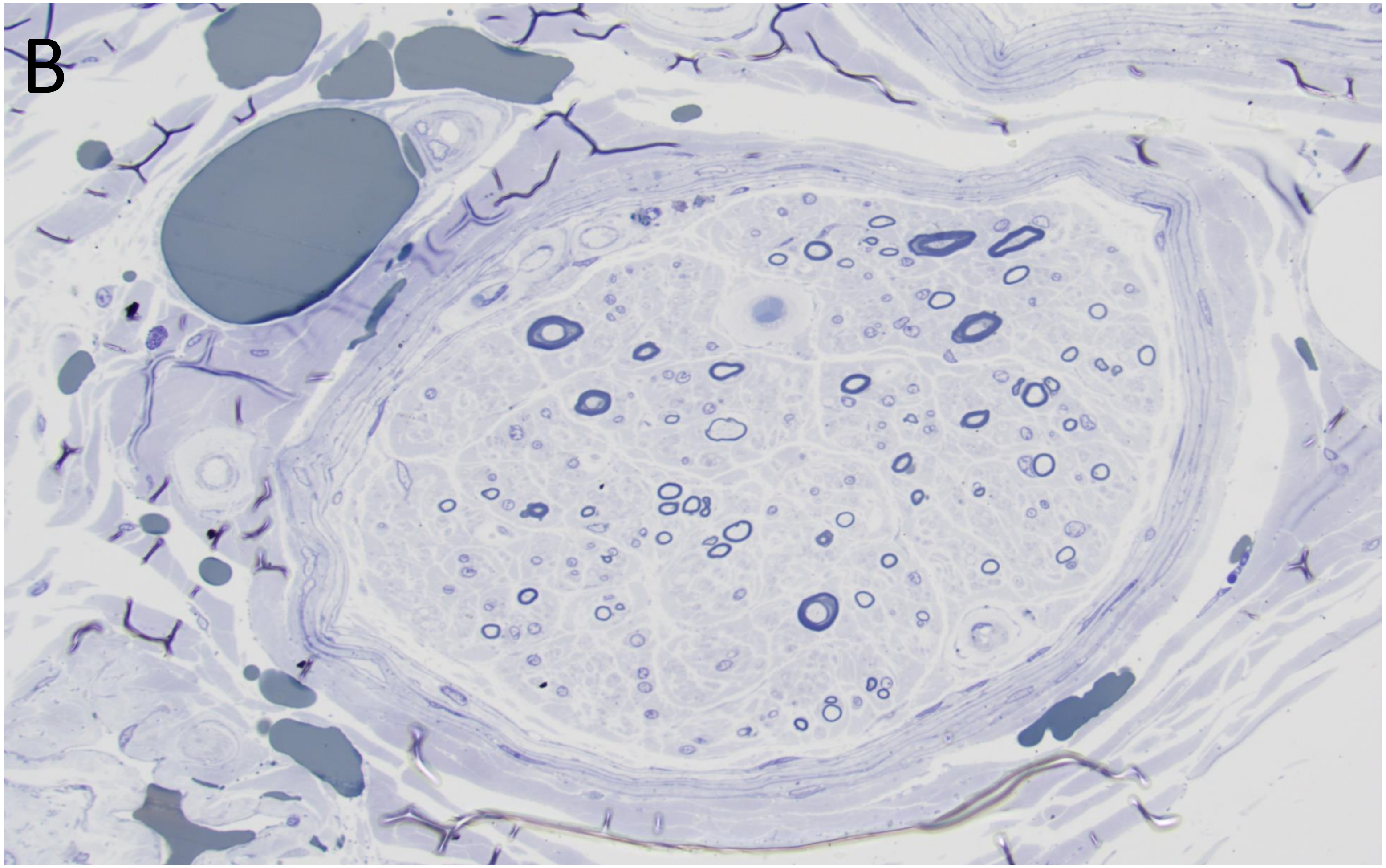
B

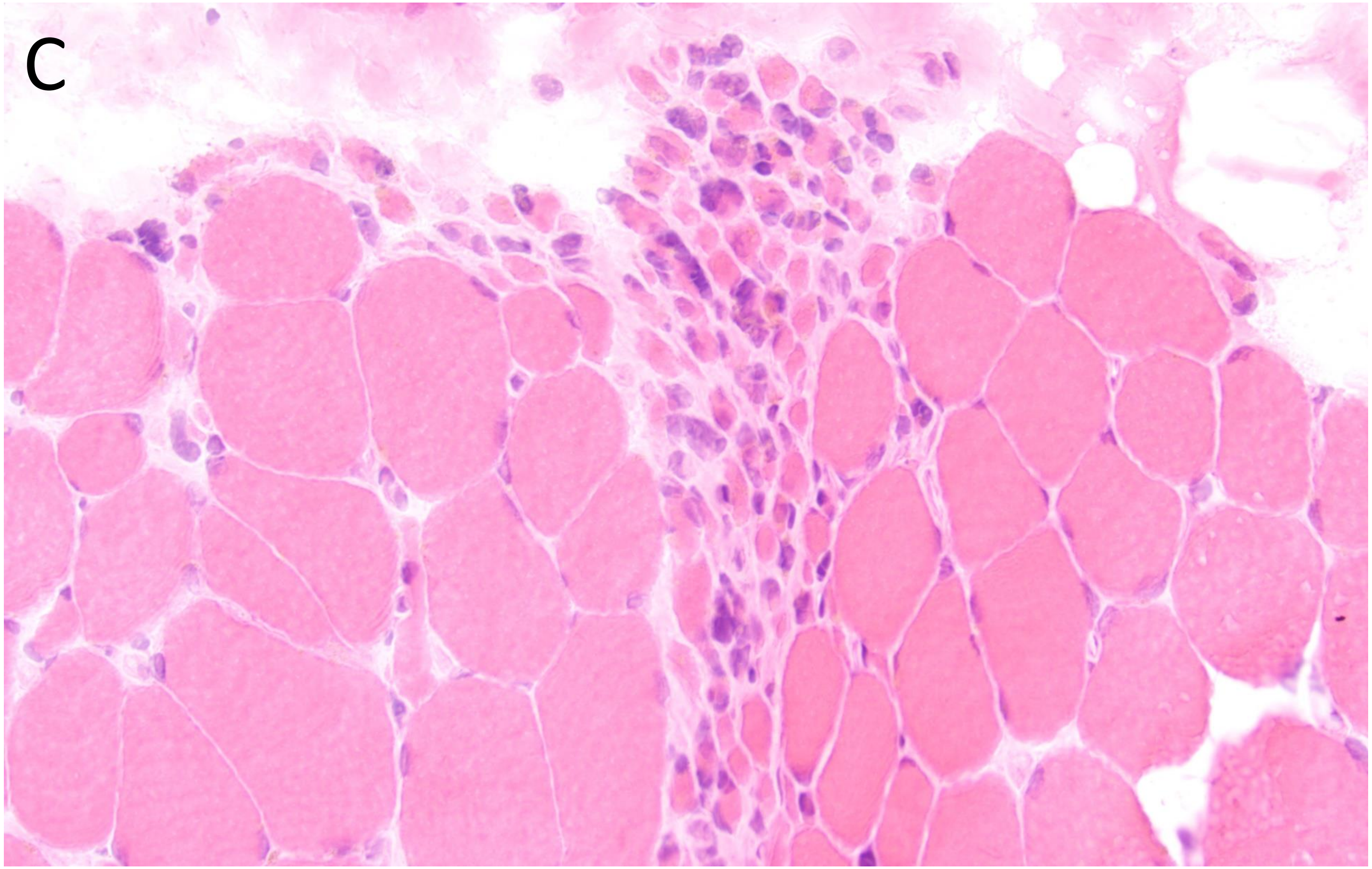
C

**Figure e1.** (A) Medulla oblongata with the inferior olive nucleus. Intranuclear p62-positive inclusions. (B) A fascicle of the sural nerve with prominent loss of myelinated fibers. Toluidine blue stained Epon section. (C) Frozen section of the skeletal muscle showing groups of atrophic fibers. Htx-eosin.

**Neurophysiological studies**

Syncope upon tilt test was present in one patient (IV:1), the response in the other 3 examined patients (IV:3, IV:6 and V: 5) was of a dysautonomic type (3) (Table 2).

**2. Confirmation of linkage on chromosome 16**

Multipoint linkage analysis confirmed the linkage with a Maximum LOD score of 3.7 on chromosome 16. Linkage peak is located in a 3.69 cM region between D16S415 and D16S515 (Figure 2).

**Biochemistry**

Two patients from family 1 (IV:1 and IV:3) went through lumbar puncture. Standard parameters were normal in both cases but in the index case of this family some oligoclonal bands were found. Furthermore, we found a mild reduction of CSF β-amyloid and mild elevation of CSF-tau in the index case. In both cases normal levels of 5-hydroxyindoleacetic acid (5-HIAA) but elevated homovanillic acid (HVA) were found. Thus, the ratio HVA/5-HIAA was elevated indicating impaired serotonergic metabolism. Iin addition, 4-hydroxy-3-methoxyphenyl glycol (HMPG) was reduced in both cases (Table 3). Levels of NfL, S100 and GFA-p normal were normal.

Autopsy studies on one patient (IV:3 and III:2 respectively) from each family and two matched sex-and aged controls showed that the protein levels of GAD65 is reduced in in the temporal cortex of the two SCA4 patients (Figure e2). Alpha-fetoprotein (AFP) levels in plasma were normal in three patients from each family.

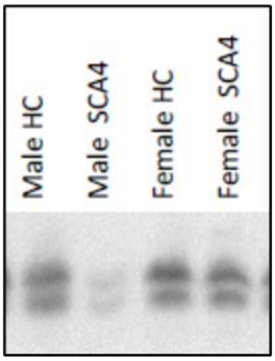


**Figure e2:** To the right temporal cortex with lower GAD65 levels in two SCA4 patients. HC: Healthy control.

**3.2 Pathology**

A right sural nerve biopsy from patient IV:1 in family 1 displayed a moderate loss of myelinated fibers mostly in small fibers. There was a suggestion of segmental demyelination. In addition, a *postmortem* sample from the left anterior tibial muscle was analysed. This sample revealed variable fiber size and fiber atrophy. The number of centrally located nuclei was not increased. The ATPase staining displayed predominance of type 1 fibers. Taken together, these findings are compatible with underlying neurogenic damage.

**4. Genetics**

One individual in family 2 (III:2) harboured an intermediate *ATXN8OS* allel on Expansion Hunter, determined to be benign. Two other individuals from the same family, one affected and one unaffected, also showed common heterozygous expansion in the *RFC1* AR disease locus.

**5. Discussion**

AO in our families (46.5) is later than in the two previous families (38.3 and 39.3 years, respectively) whereas DD is like previous descriptions (1, 2). Axonal neuropathies and neuronopathy occur in other SCAs, this trait is more prominent in SCA1, 2, 3 and 7 (16, 17). For instance, SCA1 and SCA2 individuals are more commonly affected by neuronopathy while SCA3 and SCA7 individuals were found to have both neuronopathy and axonal neuropathy (16). Neurophysiologic abnormalities are present in presymptomatic individuals affected by other SCA types, tracking these abnormalities has been proposed as a surrogate marker for SCA2 (18). The abnormal responses for SSR and RR variability are compatible with neurodegeneration of sympathetic and parasympathetic fibers.

Hearing loss was present in 22.2% of our patients; in 3 of them it can be argued they likely reflect presbyacusis. In a fourth case with asymmetric hearing loss in (II:5) the underlying could not be determined. These figures must be contrasted with SCA31, in which up to 43% of patients develop hearing loss but an at earlier age (19). Of note, SCA31 is also linked to chromosome 16 but found to be associated with nucleotide expansions in *BEAN1* (20). Furthermore, dysautonomia was found to be a rare feature of SCA31 (19).

Flanigan *et al* suggested SCA4 bore resemblance with Biemond's ataxia described in 1954 (1), however, in Family S described by Biemond in 1954, consanguinity occurred in two consecutive generations (22), (figure e3 and table e5). Besides pseudodominance, this family displayed features unseen in SCA4 such as subacute onset, optic atrophy, blindness, deafness, chorea, lack of progression and absence of cerebellar atrophy. In addition, three of these patients suffered from starvation (case 4) or from forced work during World War II (Patients 2 and 3). Of note, Patient 2 already had subacute onset of symptoms during an airway infection and before starvation. Symptom onset in the other two exposed to starvation was also subacute. Severe nutritional deficits may thus have caused sensory ataxia and exacerbated an underlying ataxia. Strachan's syndrome is a historically well characterized condition in prisoners-of-war and among persons with reduced intake of B-group vitamins and toxic exposure to tobacco (23). The aggressive course of disease, duration of 6.5 years, in cases 1 and 6 of Family S, resembles adult-onset peroxisome biogenesis disorder (24) or a mitochondrial ataxia (25). The remaining cases in pedigree S did not progress. Biemond described degeneration of the posterior columns, trigeminus nucleus, notorious demyelination and mild loss of PC (20) but not loss of motor neurons as we have demonstrated in two SCA4 cases. Hellenbroich *et al* found damage to the trigeminus nucleus but we did not (26). On the other hand, and similar to what Biemond described we found degeneration of the posterior columns. Two more reports described as Biemond's ataxia have been published since 1954. The first of these descriptions was a two-generation's Indian kindred. None of the parents exhibited neurological symptoms, all the three sons in this family presented symptoms during the first decade of life, their phenotype consisted of biopsy-proven demyelinating neuropathy, scoliosis and camptodactily (27). Biemond emphasized the lack of scoliosis and malformations in family S, therefore the disease described by Singh et al seems to be more compatible with an autosomal recessive disorder such as posterior column ataxia with retinitis pigmentosa (AXPC1) or a similar condition (28). Later, an Italian-American family with neuropathological features very similar to those described by A. Biemond was reported (29). The index case and his sisters developed late-onset ataxia but none of the parents were affected. Linkage analysis was not performed. Taken together, we provide arguments to reasonably refute SCA4 as a *forme fruste* for Biemond's ataxia.  The exact etiology of Biemond's ataxia remains unknown but some of the patients in family S were likely affected by Strachan's syndrome and two others by a condition in the spectrum of peroxisome biogenesis disorders or a mitochondrial ataxia syndrome.

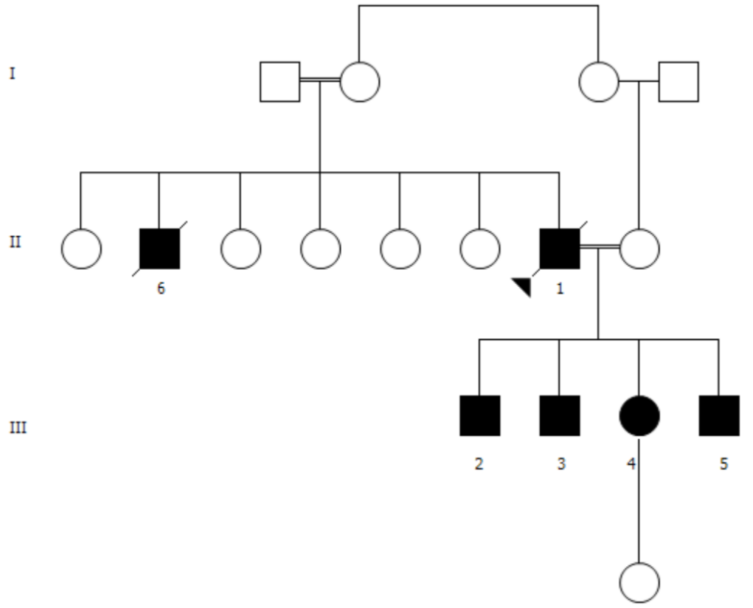


**Figure e3.** Pedigree  of family S reported by Ariel Biemond in 1954. Of note is the ocurrence of consnaguinity in two consecutive generations and also the exposure to starvation during World War II.

Zink finger homeobox 3 (*ZFHX3*), also known as AT motif binding factor 1 (*ATBF1*), cDNA was first isolated from human hepatoma cells, and its product found to bind to an AT-rich enhancer element of the human a-fetoprotein gene (*AFP*) (29). Of interest, *ATBF1* was also found to downregulate AFP levels; of note, increased AFP levels are a biomarker for ataxia-telangiectasia (A-T) (30). The *ZFHX3* promotor is activated by ATM serine/threonine kinase protein (ATM) and regulates adhesion molecules (procollagen type III α1 and integrin α8) and *PDGRGB* (31). In this context, it noteworthy that we found no changes in AFP plasma levels.

There are two isoforms (ATFB1-A and B), the ATBF1-A mRNA is mainly expressed in developing mouse brain (32). The isoforms differ mainly in the 5' end, and both include the terminal coding exon with the polyglycine stretch, often interrupted by one serine (33). The pattern of expression was studied in developing rat brain; however, the spinal cord was not included in the assessment (33). The protein was most intense in at embryonic day 20 and highly expressed in the midbrain and diencephalon whereas mRNA was highly expressed in in the brainstem (during embryogenesis and in neonatal brains), the major site of expression was in postmitotic brainstem neurons (33). Expression remained also in mature neurons with DOPA decarboxylase (DCC) activity, it is expressed to a less degree in cerebellar nuclei but not in cerebellar cortex. Outside the CNS, IHC staining was found in the nuclei of immature neurons (adrenal medulla and nerve ganglion) and in pulmonary smooth muscles (33). *Zhfx3* is highly expressed in the suprachiasmatic nucleus of the hypothalamus (34), induced missense variant in *zfhx3* accelerates circadian rhythms in mice (35). Similarly, mice with a heterozygous N-ethyl-N-nitrosourea (ENU) induced null mutation barely survive to weaning, whereas its homozygous is also lethal (35).

| Chr | Position | Gene | ExonicFunc | AAChange | 1000G_ALL** | dbSNP142 |
|---|---|---|---|---|---|---|
| **16** | 71509779 | ZNF19 | nonsynonymous | NM_006961:c.G671A:p.R224Q | 0.01 | rs10500557 |
| **16** | 72110467 | HPR | synonymous | NM_020995:c.G534T:p.V178V | 0.0009 | rs675973 |
| **16** | 72132832 | DHX38 | synonymous | NM_014003:c.G771A:p.V257V | 0.0014 | rs150221434 |
| **16** | 72821618 | ZFHX3 | synonymous | NM_001164766:c.T7815C:p.G2605G | | rs112443847 |
| **16** | 72821642 | ZFHX3 | synonymous | NM_001164766:c.T7791C:p.G2597G | | rs367610293 |

*GRCh37/hg19.
**Minor allele frequency from 1000 Genome project.

**Table e1:** Rare exonic variants that segregate in family 1.

| Metabolite (Reference interval and unit) | Patient IV:1 | Patient IV:3 |
|---|---|---|
| GFA-p ( < 1250 ng/L) | 640 | 840 |
| NfL (<1850 ng/L) | 1430 | 340 |
| Tau (<400 ng/L) | 263 | 409* |
| Beta-amyloid (>450 ng/L) | 829 | 419* |
| Phospho-tau (<80 ng/L) | 40 | 68 |
| S-100 (< 1.7 µg/L) | 0.81 | 1.2 |
| HVA (40-170 nmol/L) | 221* | 305* |
| 5-HIAA (50-170 nmol/L) | 119 | 108 |
| HVA/5-HIAA (1.8-2.3) | 1.86 | 2.8* |
| HMPG (65-140 nmol/L) | 40* | 45* |

### Table e2

CSF analysis in two patients from the first SCA4 pedigree. 0005-HIAA: 5-hydroxyindoleacetic acid, HMPG: 4-hydroxy-3-methoxyphenyl glycol, HVA: homovanillic acid. * Indicates abnormal value. Abnormal HVA/5-HIAA ratio in case IV:3 indicates reduced serotonergic metabolism. The index case had also some oligoclonal bands. Alpha-fetoprotein (AFP) levels in plasma were normal in three patients from each SCA4 family.

| **WGS** | **ExHu ZFHX3** | **Family, patient** |
|---|---|---|
| SR | (21,**50**) | Family 1, V:4 |
| SR | (21, **64**) | Family 1, IV:8 |
| SR | (21, **46**) | Family 1, IV:4 |
| SR | (21, **47**) | Family 2. III:2 |
| SR | (21,**53**) | Family 2, IV:4 |
| SR, LR | (21,21) | Family 2, III:3 |
| SR, LR | (21, **57**) | Family 3, III:2 |

**Table e3:** Locus sizing with Expansion Hunter (ExHu) v4.0.1 hg19 detects heterozygous expanded loci in *ZFHX3* for affected family members, whereas an unaffected member showed the most common normal locus configuration: homozygous for 21 copies. As with other Expansion Hunter detected loci, the sizing for larger than read length expansions is not to be interpreted as an exact size, but as flagged pathologically expanded.

| | SCA | Control |
|---|---|---|
| mesencephalon | 0.33±0.01% | 0.35±0.03% |
| pons | 1.20±0.13% | 1.58±0.12% |
| medulla oblongata | 0.35±0.05% | 0.50±0.04% |
| cerebellum | 9.25±0.68% | 11.48±0.47% |
| spinal cord | 5.5±0.4 mm | 6.7±0.3 mm |
| peripheral nerve max FA | 0.45±0.01 | 0.57±0.04 |
| peripheral nerve MTR | 14±7% | 18±3% |
| muscle MTR | 39±1% | 40±1% |

| Patient/Control | +Gender |
|---|---|
| #1 | male/male |
| #2 | female/female |
| #3 | male/male |
| #4 | male/male |

**Table e4:** Quantification of MRI examinations

| **Parameter/Feature** | **Biemond's ataxia** | **SCA4** |
|---|---|---|
| Age of onset (years) | 17-46 y | 20-60 y |
| Presentation | Subacute in 3 of 6 patients | Insidious |
| Progressive pattern | No[a] | Yes |
| Pattern of inheritance | Consanguinity in two consecutive generations | Autosomal dominant |
| Cerebellar ataxia | Yes | Yes |
| Sensory ataxia | Yes | Yes |
| Cerebellar atrophy | No[b] | Yes |
| Neuropathology | Degeneration of posterior columns<br>Neuronal oss in the trigeminal nucleus<br>Mild loss of Purkinje cells | Widespread gliosis and loss of lower motor neurons.<br>Neuronal loss in brainstem nuclei<br>Mild-moderate loss of Purkinje cells |
| Areflexia | Yes | Yes |
| Dysautonomia | No | Yes |
| Modulating factors | Starvation (in 3 patients) | Absent |
| Optic atrophy | Yes (in 2 patients) | No |
| Hearing loss | Deafness in one | Presbycusis in 4 patients |

**Table e5:** Consanguinity suggests rather and autosomal recessive pattern in family S described by A. Biemond. [a] Only one of the patients had a clear progressive character, another one improved, whereas the remaining four patients had a non-progressive course. [b] Neuropathological assessment was done in 1 case.
